# A stochastic model for COVID-19 spread and the effects of Alert Level 4 in Aotearoa New Zealand

**DOI:** 10.1101/2020.04.08.20058743

**Authors:** Michael J. Plank, Rachelle N. Binny, Shaun C. Hendy, Audrey Lustig, Alex James, Nicholas Steyn

## Abstract

While case numbers remain low, population-wide control methods combined with efficient tracing, testing, and case isolation, offer the opportunity for New Zealand to contain and eliminate COVID-19. We use a stochastic model to investigate containment and elimination scenarios for COVID-19 in New Zealand, as the country considers the exit from its four week period of strong Level 4 population-wide control measures. In particular we consider how the effectiveness of its case isolation operations influence the outcome of lifting these strong population-wide controls. The model is parameterised for New Zealand and is initialised using current case data, although we do not make use of information concerning the geographic dispersion of cases and the model is not stratified for age or co-morbidities.

We find that fast tracing and case isolation (i.e. operations that are sustained at rates comparable to that at the early stages of New Zealand’s response) can lead to containment or elimination, as long as strong population-wide controls remain in place. Slow case isolation can lead to containment (but not elimination) as long as strong Level 4 population-wide controls remain in place. However, we find that relaxing strong population-wide controls after four weeks will most likely lead to a further outbreak, although the speed of growth of this outbreak can be reduced by fast case isolation, by tracing, testing, or otherwise. We find that elimination is only likely if case isolation is combined with strong population-wide controls that are maintained for longer than four weeks.

Further versions of this model will include an age-structured population as well as considering the effects of geographic dispersion and contact network structure, the possibility of regional containment combined with inter-regional travel restrictions, and the potential for harm to at risk communities and essential workers.

**Executive Summary:**

- While New Zealand case numbers remain low, tracing, testing, and rapid case isolation, combined with population-wide control methods, offer an opportunity for the country to contain and eliminate COVID-19.
- Simulations using our model suggest that the current population-wide controls (Alert Level 4) have already had a significant effect on new case numbers (see figure below).

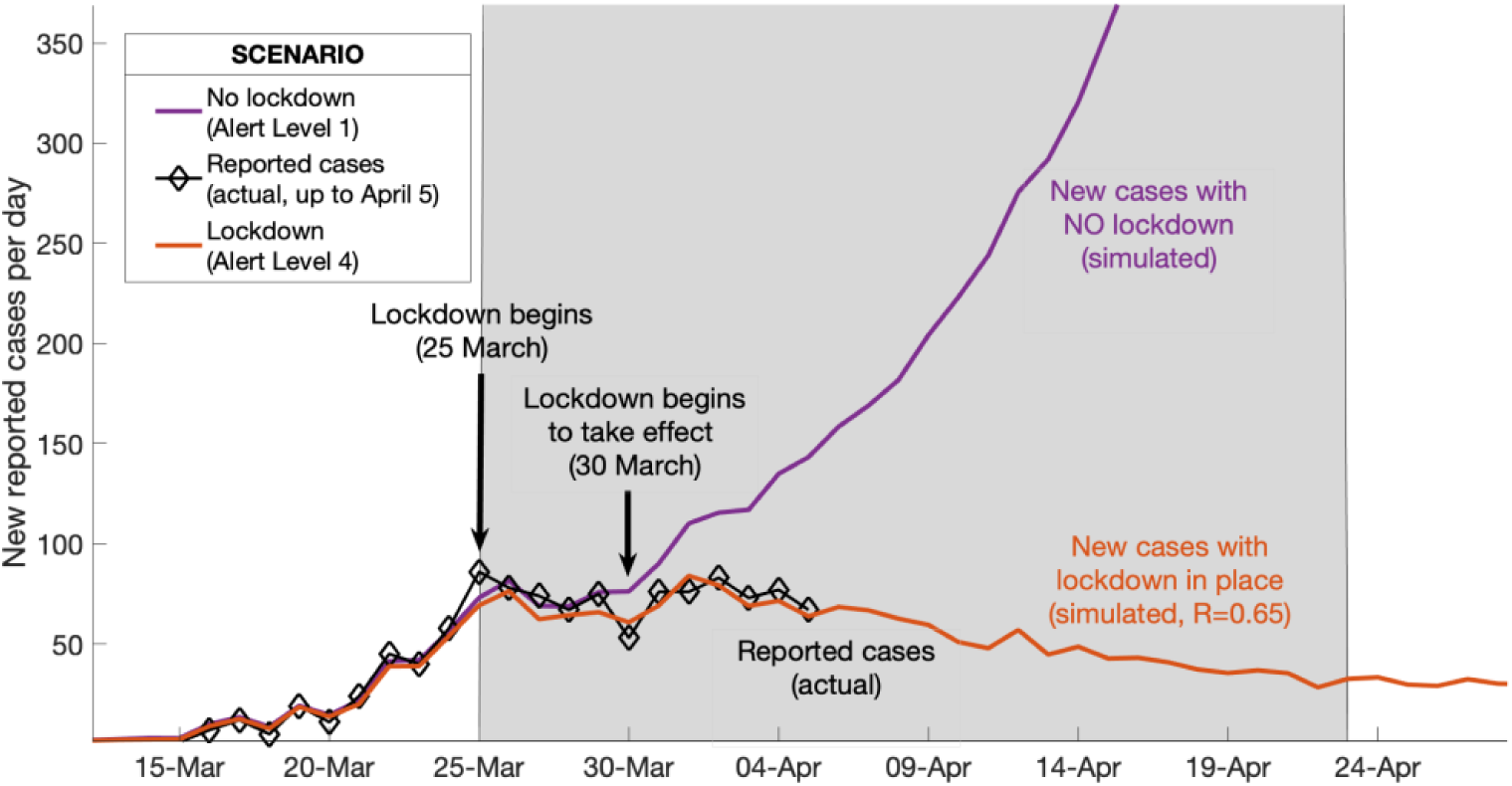
- We also find that fast case isolation, whether as a result of contact tracing, rapid testing, or otherwise, can lead to containment and possibly even elimination, when combined with strong population-wide controls.
- Slow case isolation can also lead to containment, but only as long as strong population wide controls remain in place. It is unlikely to lead to elimination.

## Introduction

The COVID-19 outbreak originated in Wuhan China in November 2019 (WHO, 28 February 2020) before spreading globally to become a pandemic in March 2020 (WHO, 11 March 2020). The human population currently lacks immunity to COVID-19, a viral zoonotic disease with reported fatality rates that are of the order of 1% (Verity et al 2020). Many countries have experienced community transmission after undetected introductions of the disease by travellers exposed in China. This has led to exponential growth of new infections in many countries, even as China, through the use of strong controls and rapid testing and tracing, has apparently managed to eliminate the disease.

At the time of writing (8 April 2020) New Zealand has yet to see significant community transmission with the majority of its detected cases directly or indirectly related to international travel (Ministry of Health, 2020). After imposing strong border restrictions in mid-March and very strong population-wide control measures in late March, the number of new cases detected per day has plateaued, presenting the country with an opportunity to explore strategies that are not available to other countries. While case numbers remain small (through the imposition of population-wide controls or otherwise), tracing, and case isolation operations may be able to remain ahead of infection rates, resulting in containment, and opening up the possibility for elimination.

A number of deterministic compartment models have been developed for the spread of COVID-19 internationally (e.g. Ferguson et al 2020), as well several that have been specifically adapted for New Zealand (James et al 2020, Wilson 2020). Such models are expected to describe the progression of an epidemic after the onset of community transmission and at high population densities where random encounters are important sources of infection. They have been used to explore long-term consequences and strategies for New Zealand should the disease not be contained, but at the early stages of an outbreak or in regions with low population densities, stochastic models are expected to be more appropriate. In particular, stochastic models can be used to explore scenarios with tracing and case isolation or alternative testing strategies which may lead to containment or even elimination of the disease.

In this study, we introduce a continuous-time branching process model similar to that of Davies et al (Davies 2020) (although our model is currently not age-structured) and explore different scenarios for case isolation and population-wide control interventions. In particular we use the model to explore the possible scenarios if New Zealand were to exit its current highly restrictive control measures in late April, after four weeks. The model is intended for use only when case numbers are small relative to population size and herd immunity is not prevalent in limiting secondary cases. It may, however, provide insight into short-term scenarios that explore New Zealand’s options towards the end of its first four-week Alert Level 4 lockdown period. Longer term scenarios can be considered using the model if herd immunity is added to the model, but other models should also be considered in these circumstances.

## Methods

We use a continuous-time branching process to model the number of infections, with initial seed cases representing overseas arrivals. Key model assumptions are:

- Infected individuals are grouped into two categories: (i) those who show clinical symptoms at some point during their infection; and (ii) those who are subclinical. Each new infection is randomly assigned as subclinical with probability *p*_sub_ = 0.33 and clinical with probability 1 – *p*_*sub*_, independent of who infected them. Once assigned as clinical or subclinical, individuals remain in this category for the duration of their infectious period.
- In the absence of self-isolation measures (see below), each infected individual *i* causes a randomly generated number *N*_*i*_*∼Poisson*(*R*_*i*_) of new infections. For clinical individuals, *R*_*i*_ = *R*_*clin*_ and for subclinical individuals, *R*_*i*_ = *R*_*sub*_, which is assumed to be 50% of *R*_*clin*_ (Davies 2020).
- The model can be generalised to include additional sources of individual heterogeneity by drawing *R*_*i*_ is drawn from a specified distribution (e.g. Lloyd-Smith 2005), instead of simply one of two fixed values *R*_*clin*_ and *R*_*sub*_. However for simplicity we ignore individual heterogeneity within the clinical and subclinical categories.
- The time between an individual becoming infected and infecting another individual, the generation time *T*_*G*_, follows a Weibull distribution (*W*) with mean and median equal to 5.0 days and standard deviation of 1.9 days (Feretti 2020). The infection times of all *N*_*i*_ secondary infections from individual *i* are independent, identically distributed random variables from this distribution (see Figure 1).
- Clinical individuals have an initial period during which they are either asymptomatic or have sufficiently mild symptoms that they have not self-isolated. During this period, their infectiousness is as shown by the blue curve in Figure 1. At the end of this period, once they have developed more serious symptoms, they are isolated and their infectiousness reduces to *c*_*iso*_ = 65% (Davies 2020) of the value it would have without isolation (green curve in Figure 1). This represents a control policy of requiring symptomatic individuals to self-isolate.
- Subclinical individuals do not get isolated and are not reported in case data.
- All individuals are assumed to be no longer infectious 30 days after being infected. This is an upper limit for computational convenience; in practice, individuals have very low infectiousness after about 12 days because of the shape of the generation time distribution (Fig. 1).
- Individuals who have recovered from the virus are assumed to have immunity for the duration of the period simulated and cannot be infected again. This means that the proportion of the population that is susceptible at time *t* is 1 – *N*(*t*)/*N*_*pop*_, where *N*(*t*) is the cumulative number of infections at time *t* and *N*_*pop*_ is the total population size.
- The time *T*_*iso*_ between infection and isolation the sum of two random variables *T*_1_ and *T*_2_. *T*_1_ represents the incubation period (time from infection to onset of symptoms) and has a gamma distribution with mean 5.5 days and shape parameter 5.8 (Lauer 2020). *T*_2_ represents the time from onset to isolation and is taken from New Zealand case data (see below).
- The model does not explicitly include a latent period or pre-symptomatic period. However, the shape of the Weibull generation time distribution (Figure 1) captures these phases, giving a low probability of a short generation time between infections and with 90% of infections occurring between 2.0 days and 8.4 days after infection.
- The model is simulated using a time step of *δt* = 1 day. At each step, infectious individual *i* produces a Poisson distributed number of secondary infections with mean

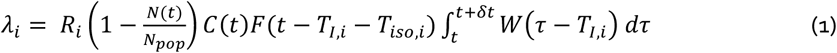

where *R*_*i*_ ∈ {*R*_*clin*_, *R*_*sub*_} is the individual’s mean number of secondary infections, *T*_*I,i*_ is time individual *i* became infected, *T*_*iso,i*_ is the delay from becoming infected to being isolated, *C*(*t*) is the control effectivity at time *t* (see below), and *F*(*t*) is a function describing the reduction in infectiousness due to isolation:

**Figure 1.**
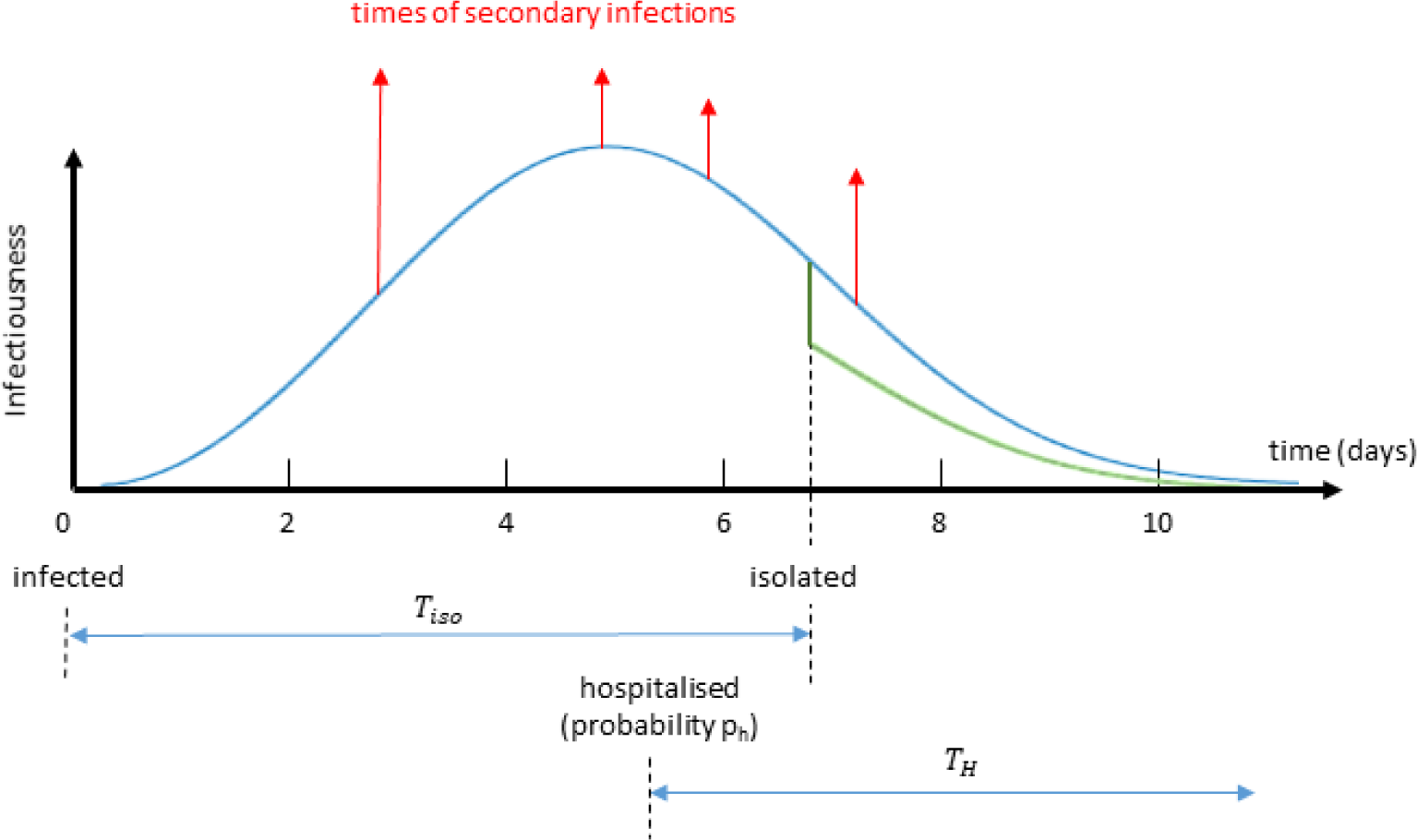
Schematic diagram showing the timeline and infection intensity of a typical infection. In the early stages secondary infections are unlikely. Red arrows show the exposure times of new secondary infections. After isolation the infection intensity is reduced to a lower level (green curve). Subclinical infectives are not isolated and follow the shape of the blue curve throughout, but with a lower overall infectiousness. Time from infection to isolation is the sum of two random variables *T*_*iso*_ = *T*_1_ + *T*_2_ drawn from shown in Table 1. Clinical infections have probability *p*_*h*_ of hospitalisation; length of hospital stay *T*_*H*_ is a random variable drawn from the distribution shown in Table 1.

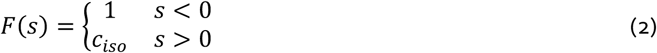

- Clinical infections have a probability *p*_*H*_ = 7.8% of being hospitalised, equivalent to an overall infection-hospitalisation ratio of 5.25% (Verity et al 2020). The duration of hospital stay is exponentially distributed with mean 10 days (Zhou et al, 2020). For simplicity, hospitalisation is assumed to occur at the same time as onset of symptoms, i.e. time *T*_1_ after infection.
- The model was initialised with seed cases representing arrival of infected individuals from overseas. The number and timing of these seed cases was chosen to replicate real case data (see below for details).

### Case data

Data was obtained from the Ministry of Health showed *N* = 1214 confirmed and probable cases of COVID-19 in New Zealand up to 7 April 2020. For each case, the dataset contained the following fields: whether there was recent international travel history and if so date of arrival to New Zealand; date of onset of symptoms (from patient recall, where available); date of isolation.

**Table 1.**
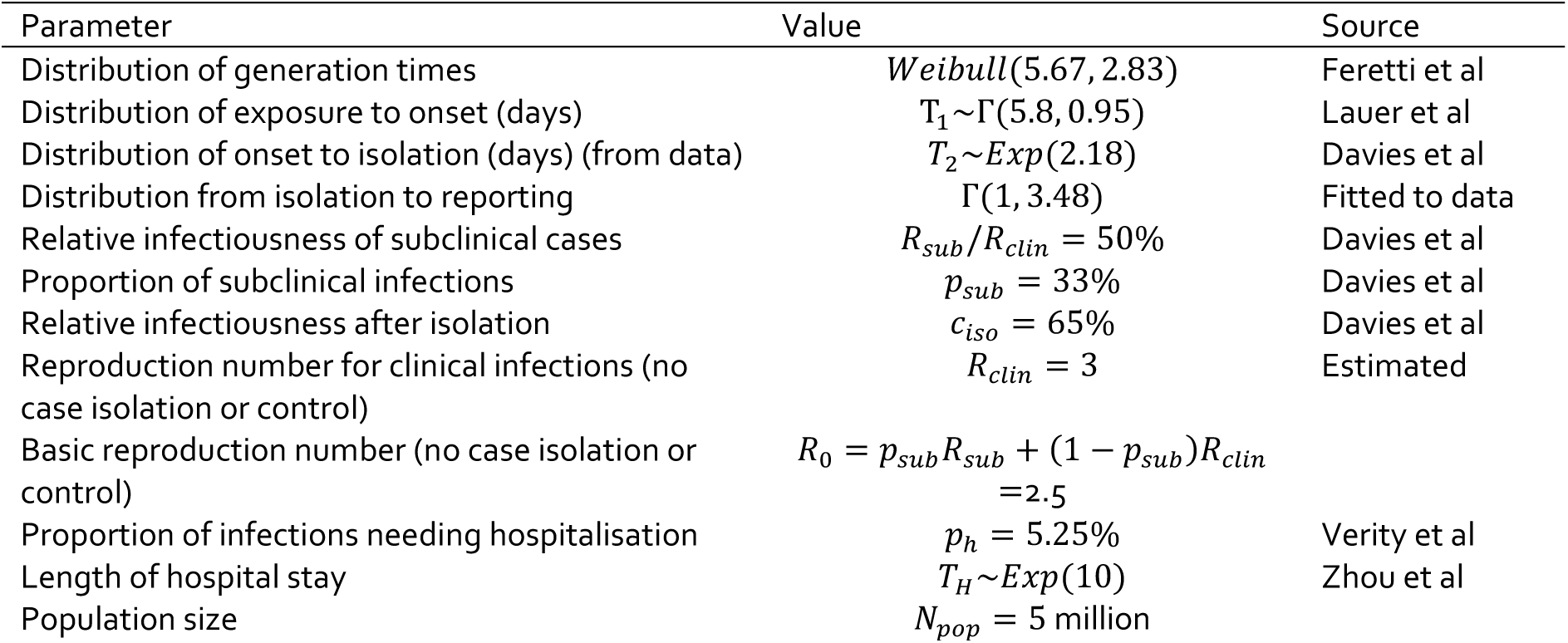
The parameters used in the model and their source.

| Parameter | Value | Source |
| --- | --- | --- |
| Distribution of generation times | $Weibull(5.67, 2.83)$ | Feretti et al |
| Distribution of exposure to onset (days) | $T_1 \sim \Gamma(5.8, 0.95)$ | Lauer et al |
| Distribution of onset to isolation (days) (from data) | $T_2 \sim Exp(2.18)$ | Davies et al |
| Distribution from isolation to reporting | $\Gamma(1, 3.48)$ | Fitted to data |
| Relative infectiousness of subclinical cases | $R_{sub}/R_{clin} = 50\%$ | Davies et al |
| Proportion of subclinical infections | $p_{sub} = 33\%$ | Davies et al |
| Relative infectiousness after isolation | $c_{iso} = 65\%$ | Davies et al |
| Reproduction number for clinical infections (no case isolation or control) | $R_{clin} = 3$ | Estimated |
| Basic reproduction number (no case isolation or control) | $R_0 = p_{sub}R_{sub} + (1 - p_{sub})R_{clin} = 2.5$ | |
| Proportion of infections needing hospitalisation | $p_h = 5.25\%$ | Verity et al |
| Length of hospital stay | $T_H \sim Exp(10)$ | Zhou et al |
| Population size | $N_{pop} = 5 \text{ million}$ | |

Model simulations were seeded with the *N*_*int*_ = 501 cases that had a known international arrival date. For these cases, the infection date was estimated backwards from the date of onset of symptoms (distribution shown in Table 1). For cases that did not include an onset date, the infection date was backdated from the arrival date. Cases missing an isolation date were assumed to remain fully infectious for the whole infectious period. Secondary infections that occurred before arrival in New Zealand were ignored. Cases that were flagged as associated with international travel but missing an arrival date were assumed to have arrived at the same time as infection, so all their secondary infections were included. To allow for the fact that the case data only includes clinical cases, an additional number *N*_*int,sub*_*∼Poisson*(*N*_*int*_*p*_*sub*_/(1 – *p*_*sub*_)) of subclinical seed cases were added. The arrival, onset and isolation dates for these subclinical seed cases were approximated by random sampling with replacement from the clinical seed cases.

The distribution of time between symptom onset and isolation date, *T*_2_, was estimated to be an exponential distribution (Kucharski 2020). The mean of the distribution, *E*(*T*_2_) = 2.18 days, was the mean taken from the *N*_*dom*_ cases that were not associated with international travel. It is likely this will be an underestimate as the data is from the early stage epidemic when there are few community transfer cases and contact tracing is more effective. Results are also shown using the more conservative estimate of *E*(*T*_2_) = 6 days (Davies 2020). We refer to these instances as *fast* and *slow* case isolation.

To allow for the time lag from isolation to reporting, each isolated case was assigned a time for the delay to reporting drawn from a gamma distribution, with parameters approximated from the *N*_*dom*_ domestic cases (see Table 1). The international cases were assigned the actual reporting date as recorded in the data.

### Control sub-model

Population-wide control interventions were modelled via the function *C*(*t*), which represents the transmission rate relative to no population-wide control. Population-wide control interventions include closure of schools, universities or non-essential businesses, restrictions on large gatherings and domestic travel, social distancing measures and stay-at-home orders. New Zealand’s control measures are based on a scale of Alert Levels from 1 to 4. Alert Level 4 was introduced on 25 March 2020. It is currently too early to assess the effectiveness of this control so different scenarios are shown below.

### Model outputs

For each simulation, the model outputs the number of newly infected cases each day, the number of cases reported each day, and the total number of cases currently in hospital. The main focus of this paper is the short-term effects of different control interventions, so we do not show hospitalisations in most of our results. However, the model can be used to assess demand for hospital beds and fatality rates over the longer term, comparable to James et al (2020). Results show 50 realisations of the stochastic model, with the average overlaid.

### Scenarios

We consider a number of scenarios that draw on optimistic and pessimistic views on the effectiveness of New Zealand’s Alert Level system^1^. At the time of writing, New Zealand is at Alert Level 4, which includes strong population-wide controls such as:

- requiring anyone not involved in essential work to stay at home;
- the closure of educational facilities;
- the closure of businesses, except for essential services such as supermarkets, pharmacies and clinics, and lifeline utilities.

These measures were enacted on 25 March, and at the time of writing, there has been an insufficient period of time to fully observe their effectiveness from clinical case data alone. New Zealand only spent two days at Alert Level 3, and these days were unlikely to be representative of how these control measures may work in the longer term. Nonetheless, we can observe the effects of similar population-wide controls in other countries, some of which have been in place for significantly longer. We categorise these controls in two ways: by their apparent strength compared to the New Zealand Alert Level system, and by their observed effectiveness in controlling case numbers. Following Binny et al (Binny 2020), we summarise this categorisation in Table 2. It is evident from the table that the effectiveness of controls varies widely, which likely reflects the time over which the controls have been in place as well as political, social, and cultural considerations.

**Table 2.** Observed values of the effective reproduction number *R*_*eff*_ sourced from international reporting of case numbers and information concerning the levels of controls in place (Binny 2020). Exemplar countries are noted in the table for each category.

| Alert | Effectiveness |  |  |
| --- | --- | --- | --- |
|  | Low | Med | High |
| Level 4 | 2.1<br>(e.g. GBR) | 1.3-1.6 (e.g. DEU) | 0.9<br>(e.g. NOR) |
| Level 3 | 1.8<br>(e.g. USA) | 1.3<br>(e.g. NLD) | 1.0-1.1<br>(e.g. NSW) |
| Level 2 |  | 1.6-1.8 (e.g. SWE) | 1.1 (e.g. HKG) |

It is apparent that highly effective Alert Level 4-strength controls can lead to effective reproductive numbers of less than one (e.g. Denmark). However, we also observe Level-4 strength controls that are less effective, although in general we observe that in most countries that have imposed controls have found their effectiveness improves over time. New South Wales has also demonstrated highly effective Alert Level 3 strength controls, with a basic reproduction number observed to be just over one. In countries with relatively small numbers of cases (e.g. Hong Kong), Alert Level 2 measures can be highly effective when combined with sufficiently rapid tracing, testing, and isolation.

In what follows we consider optimistic, pessimistic, and realistic scenarios that attempt to span reasonable estimates of the effectiveness of controls as well as the speed of case isolation. In particular, we consider scenarios where New Zealand reduces its Alert Level from 4 to 3 from 23 April. The relative transmission rate *C*(*t*) at each alert level and under optimistic, pessimistic, and realistic scenarios, is shown in Table 3. These have been chosen by comparison with other studies (Flaxman 2020) and with observed *R*_*eff*_ internationally (Binny 2020).

**Table 3.**
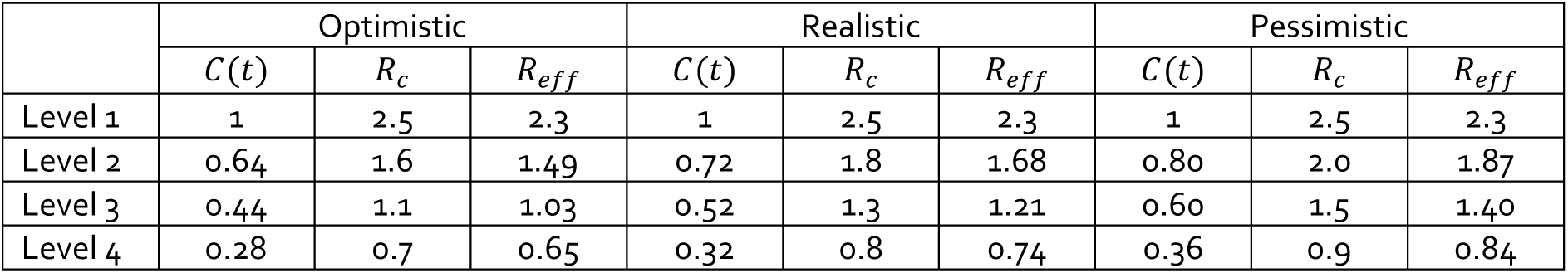
Assumed effectiveness of population-wide controls used in the model at alert levels 1-4. *C*(*t*) is the transmission rate relative to no control 1; *R*_*c*_ is the reproduction number under a given level of population-wide control, but before accounting for case isolation. *R*_*eff*_ is the effective reproduction number under a given level of population-wide control and accounting for fast case isolation (average 2.18 days from onset to isolation). Slow case isolation will give an effective reproduction number that is between the quoted values of *R*_*c*_ and *R*_*eff*_.

|  | Optimistic |  |  | Realistic |  |  | Pessimistic |  |  |
| --- | --- | --- | --- | --- | --- | --- | --- | --- | --- |
| | $C(t)$ | $R_c$ | $R_{eff}$ | $C(t)$ | $R_c$ | $R_{eff}$ | $C(t)$ | $R_c$ | $R_{eff}$ |
| Level 1 | 1 | 2.5 | 2.3 | 1 | 2.5 | 2.3 | 1 | 2.5 | 2.3 |
| Level 2 | 0.64 | 1.6 | 1.49 | 0.72 | 1.8 | 1.68 | 0.80 | 2.0 | 1.87 |
| Level 3 | 0.44 | 1.1 | 1.03 | 0.52 | 1.3 | 1.21 | 0.60 | 1.5 | 1.40 |
| Level 4 | 0.28 | 0.7 | 0.65 | 0.32 | 0.8 | 0.74 | 0.36 | 0.9 | 0.84 |

**Table 4:**
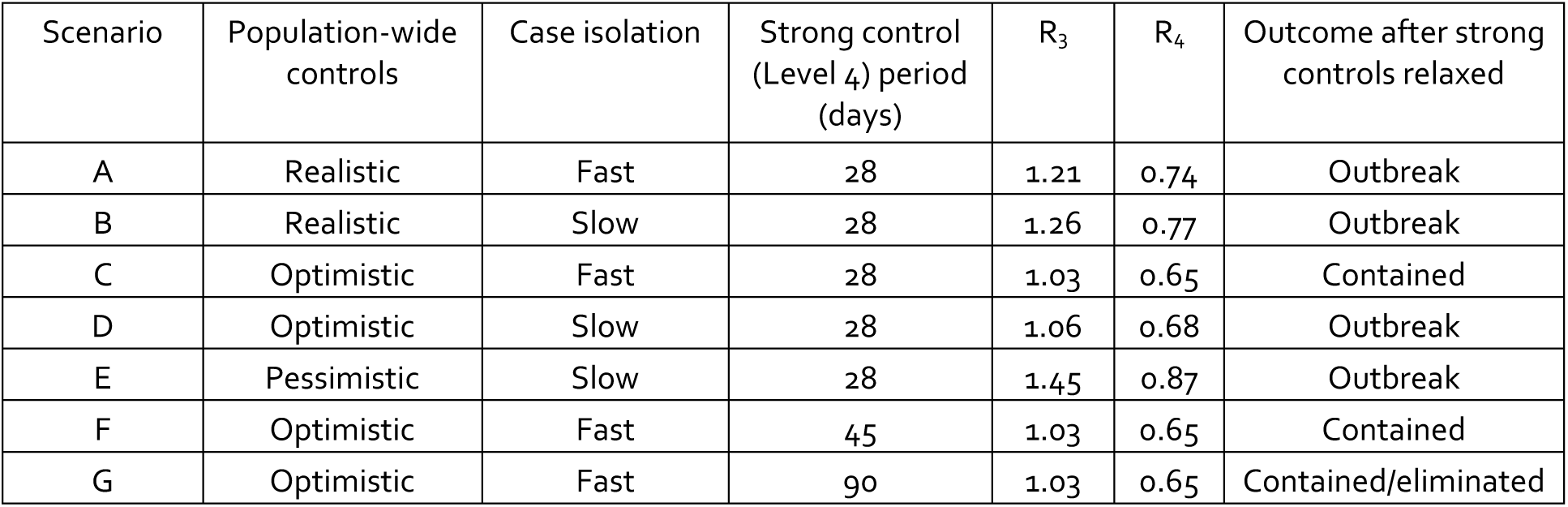
Summary of the results by scenario, showing the effective reproduction numbers during each stage (Level 4 and Level 3, R_4_ and R_3_ respectively) and the eventual outcome.

| Scenario | Population-wide controls | Case isolation | Strong control (Level 4) period (days) | $R_3$ | $R_4$ | Outcome after strong controls relaxed |
| --- | --- | --- | --- | --- | --- | --- |
| A | Realistic | Fast | 28 | 1.21 | 0.74 | Outbreak |
| B | Realistic | Slow | 28 | 1.26 | 0.77 | Outbreak |
| C | Optimistic | Fast | 28 | 1.03 | 0.65 | Contained |
| D | Optimistic | Slow | 28 | 1.06 | 0.68 | Outbreak |
| E | Pessimistic | Slow | 28 | 1.45 | 0.87 | Outbreak |
| F | Optimistic | Fast | 45 | 1.03 | 0.65 | Contained |
| G | Optimistic | Fast | 90 | 1.03 | 0.65 | Contained/eliminated |

We consider levels of effectiveness fast case isolation, by tracing or otherwise. Fast case isolation results in an expected time from the onset of symptoms to isolation of 2.18 days, as noted above. This is comparable to what occurred at an early stage in the New Zealand case data (Ministry of Health 2020), and assumes that this has been sustained despite the heavier case load. We also consider scenarios where the speed of case isolation has slowed (due to heavier case-loads for testing and contact tracing teams or otherwise), so that the expected time from symptoms to isolation has increased to 6 days (Davies 2020).

In what follows, we regard scenarios that, in 50% of stochastic realisations, eliminate all infections as having resulted in elimination. Similarly, scenarios that on average across realisations result in steady case numbers of less than 50 new cases per day are considered to be contained, while scenarios that result in unbounded growth during the simulation time as resulting in an outbreak.

## Results

### Population wide controls

We first consider the effect of the current level 4 population-wide controls that began on 25 March, relative to a scenario with only weak population-wide control (Alert Level 2) (and existing border controls). The latter provides an important counterfactual for evaluating the benefits of the current Alert Level 4 controls. We consider optimistic and pessimistic Level 4 controls applied from 25 March. These are compared to an weakly controlled (Level 2) counterfactual scenario in Figure 2, which shows *reported* cases per day in comparison with the actual data. The fact that current cases numbers have remained steady (consistently < 100 new cases per day) suggest that the lockdown has had the effect of containing case numbers. Note that the close match between model outputs and actual data up to the end of March is because the majority of cases during this period are overseas cases, which are taken directly from data.

**Figure 2:**
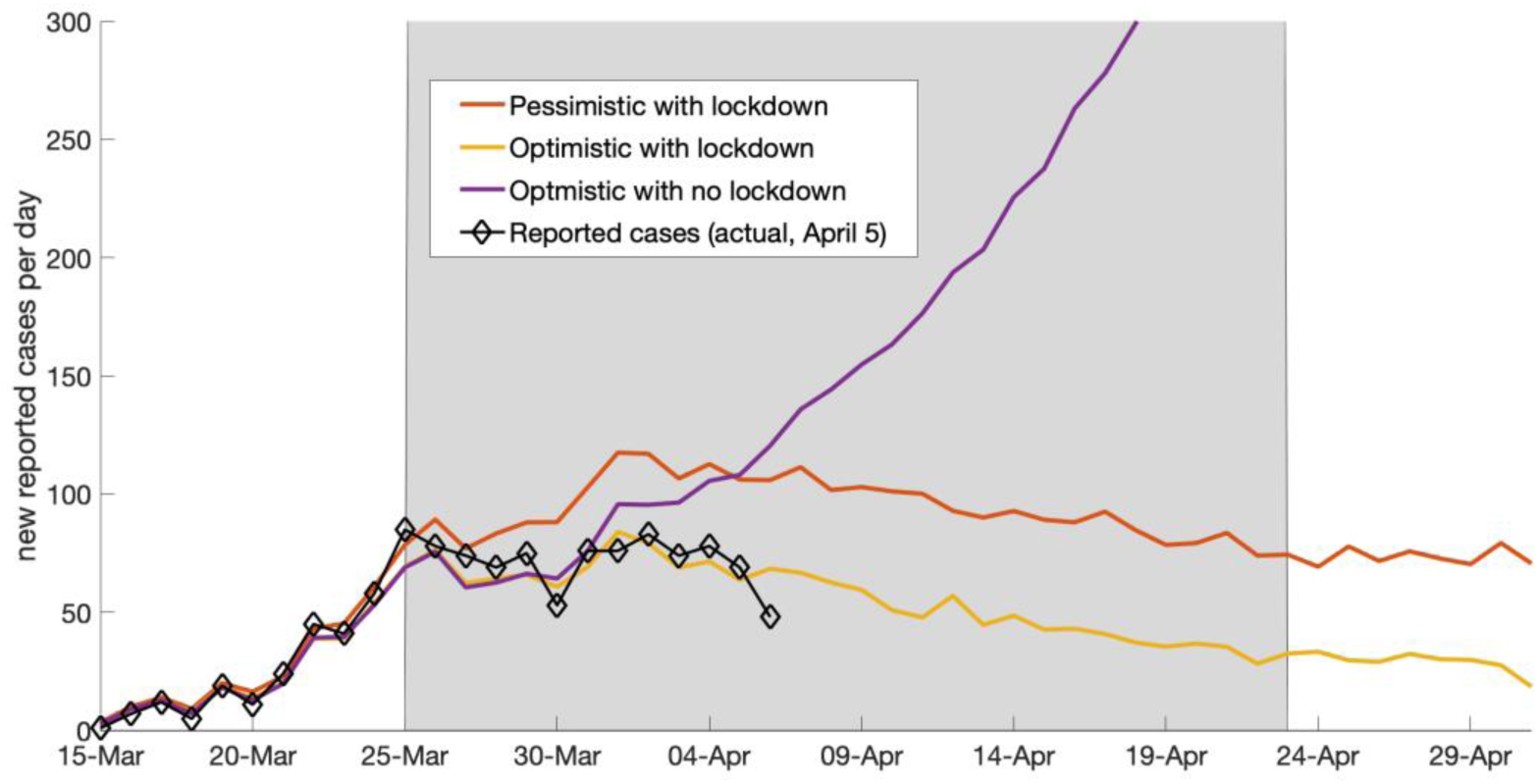
(Counterfactual scenario) Fast case isolation (expected time from symptom onset to isolation is 2.18 days) in scenarios with optimistic (red) and pessimistic (green) Alert Level 4 control effectiveness compared to a scenario with no population-wide controls (purple). For these scenarios, it was assumed that an average of 10 cases per day continue to arrive from overseas for the period simulated.

### Speed of case isolation

We also investigate the effect of the speed of case isolation in realistic control scenarios that both start with Level 2 control, i.e. only weak social distancing. On 25 March, this switches to Level 4 control. After 28 days at level 4, we switch to Level 3 control. The two scenarios differ based on the effectiveness of case isolation:

- Scenario A: a scenario where case isolation is fast. Here the expected time from the onset of symptoms to isolation is 2.2 days. This is comparable to early stage New Zealand data, and assumes that this can been sustained despite the heavier case load.
- Scenario B: a scenario where the speed of isolation have slowed so that the expected time from symptoms to isolation has dropped to 6 days.

In scenario A, the population-wide effective reproduction number *R*_*eff*_ at each alert level is as shown in Table 3. These values were calculated by simulating the number of secondary cases from a single seed case and averaging over 10^6^ realisations. Scenario A leads to a decline in daily isolated cases during Level 4 control but cases rise after switching back to level 3 (Figure 3). In scenario B, the slower case isolation scenario, the effective reproduction rates are 1.26 in level 3 and 0.77 in level 4. Now there is a slower drop during Level 4 control as cases are still rising from the previous Level 1 control period and the rise at Level 3 is faster. This is illustrated in Figure 4.

**Figure 3:**
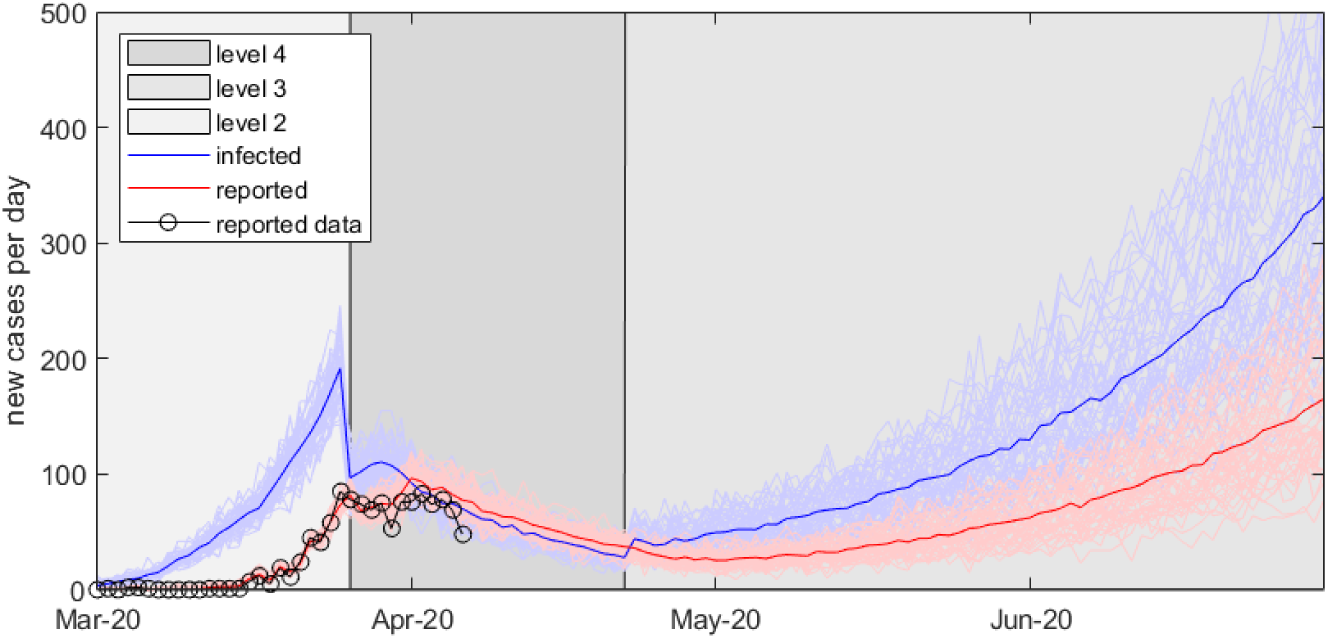
(Scenario A) Fast case isolation (expected time from symptom onset to isolation is 2.18 days) in a scenario with realistic Alert Level 4 and 3 control effectiveness. This scenario results in an outbreak.

**Figure 4:**
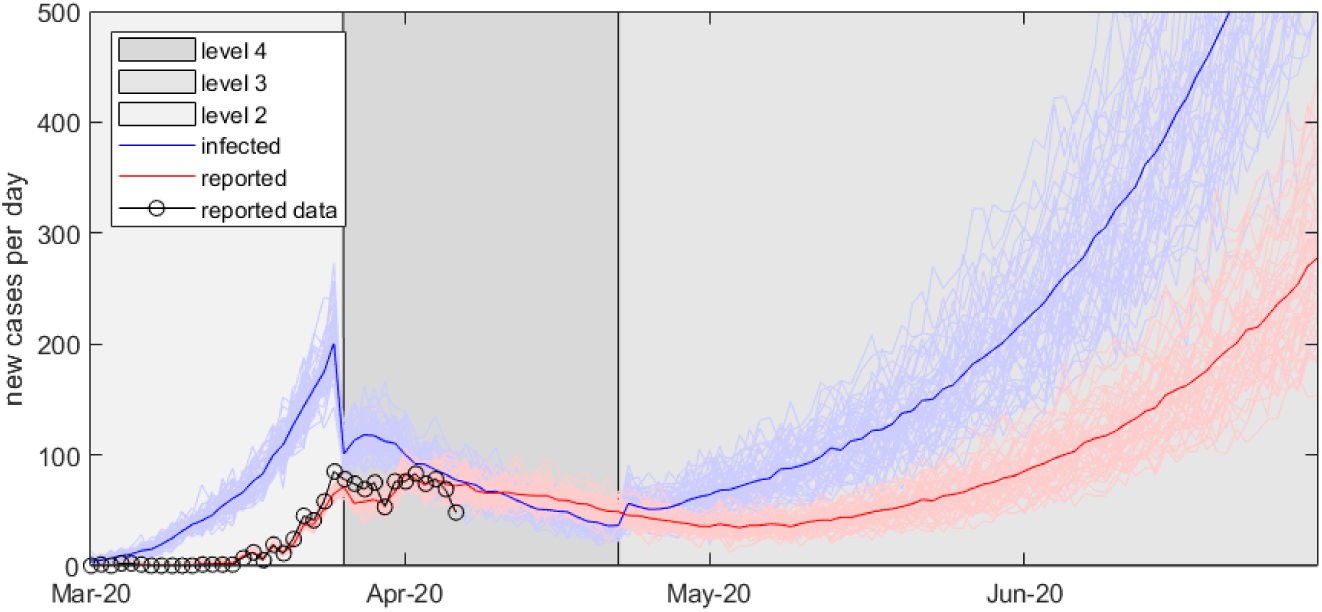
(Scenario B) Slow case isolation (expected time from symptom onset to isolation is 6 days) in a scenario with realistic Alert Level 4 and 3 control effectiveness. This scenario results in an outbreak.

An optimistic control scenario allows for the possibility of containment. Once more these scenarios begin with Level 2 control, followed by a switch to Level 4 control on 25 March. After 28 days Level 3 control is initiated. The two scenarios differ based on the speed of case isolation:

- Scenario C: case isolation remains fast (as in A).
- Scenario D: case isolation have slowed (as in B).

Figure 5 illustrates the outcome of scenario C. In this scenario, the effective reproduction numbers *R*_*eff*_ at Level 3 and Level 4 are as shown in Table 3. Case numbers are slowly reduced to a level of approximately 30 per day for the period of Level 3 controls. In scenario D however (effective reproduction numbers 1.06 and 0.68 in levels 3 and 4 respectively), cases gradually rise after the transition to Level 3 controls (not shown).

**Figure 5:**
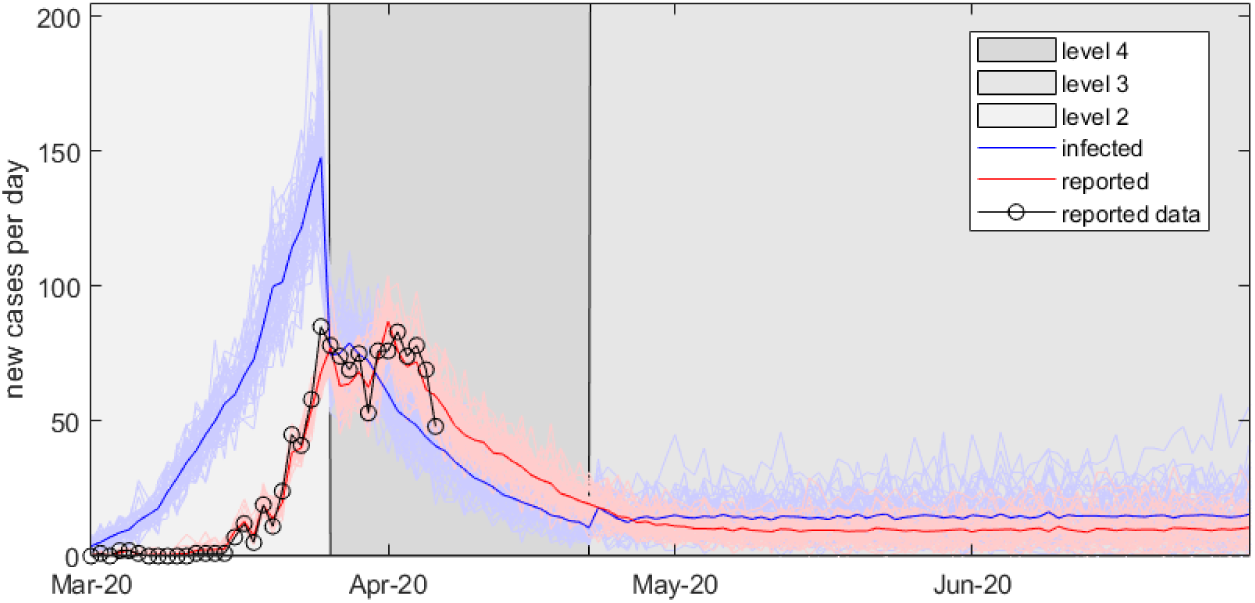
(Scenario C) Fast case isolation (expected time from symptom onset to isolation is 2.18 days) in a scenario with optimistic Alert Level 4 and 3 control effectiveness. This scenario results in containment.

Finally we consider a pessimistic scenario with slow case isolation. In this scenario (Scenario E), cases are contained during Level 4 but rebound very soon after the switch back to Level 3 (not shown). The effective reproduction numbers at Level 3 and Level 4 respectively are 1.45, and 0.87 respectively.

### Extended Level 4 controls

We also illustrate the effects of extending the Level 4 period from 28 days to either 45 days or 90 days. In an optimistic scenario with 45 days at Level 4 with fast tracing and case isolation, this leads to containment as shown in Figure 5 (Scenario F). A 90 day period at Level 4 leads to containment to very low levels by the end of the 90 day period (Scenario G), shown in Figure 6. This scenario leads to elimination in approximately 50% of stochastic realisations, although a full analysis of the probability of elimination is left for future work.

**Figure 6:**
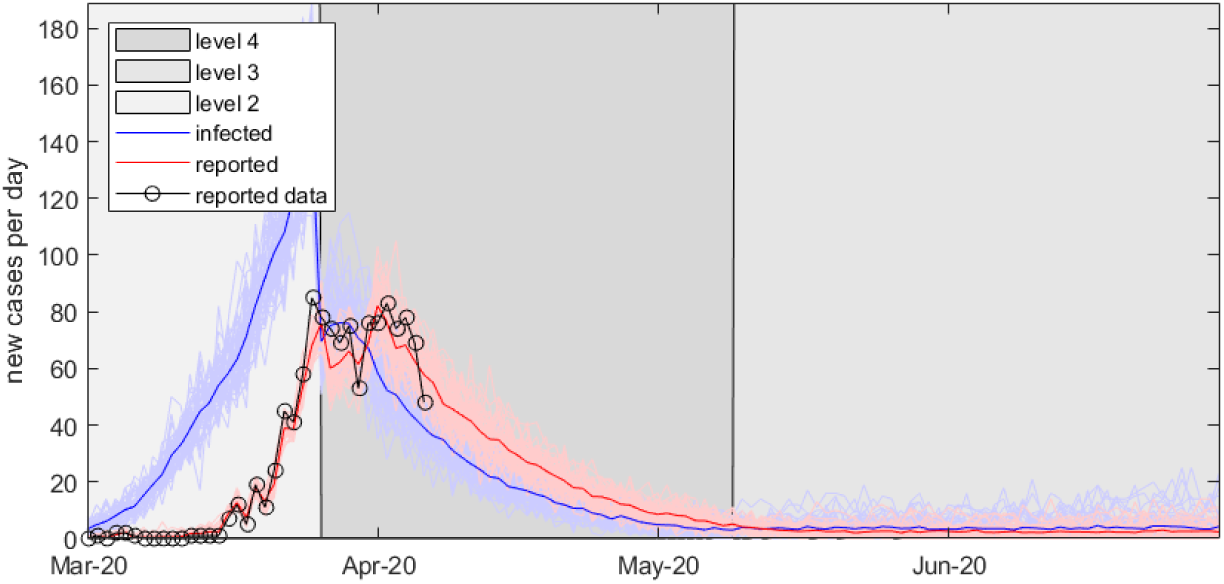
(Scenario F) Fast case isolation (expected time from symptom onset to isolation is 2 days) in a scenario where an optimistic Alert Level 4 control is extended for 45 days. This scenario results in containment.

**Figure 7:**
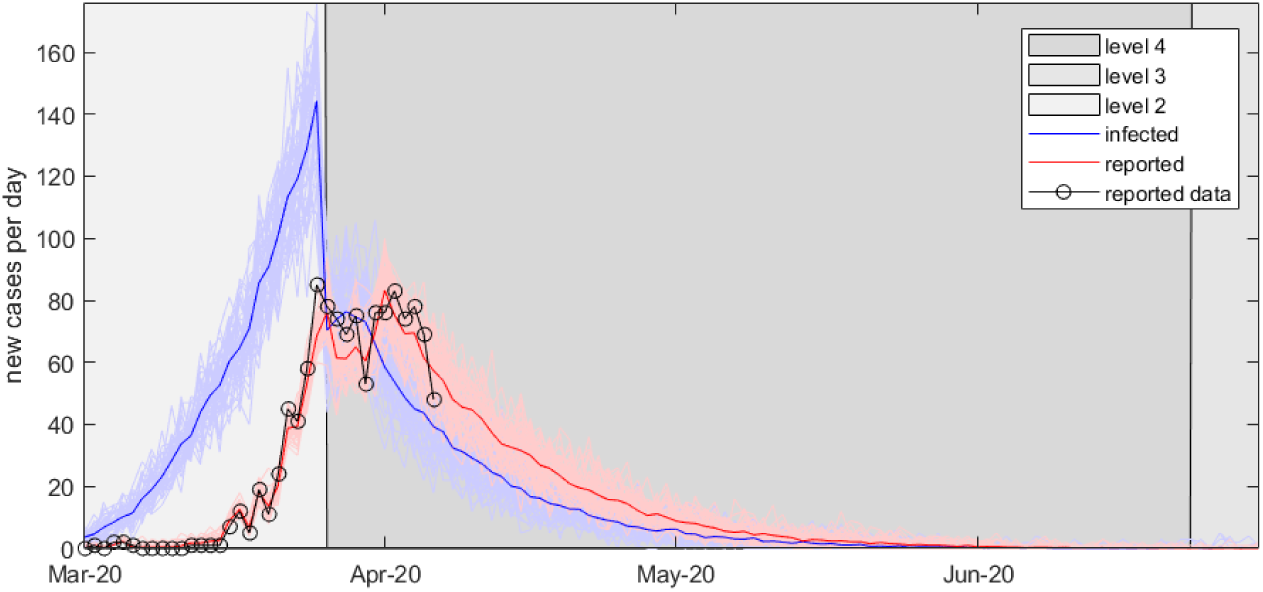
(Scenario G) Fast case isolation (expected time from symptom onset to isolation is 2 days) in a scenario where an optimistic Alert Level 4 control is extended for 90 days. This scenario results in elimination.

### Long-term simulations

Finally, although not the focus of this report, the model can also be used to simulate longer term scenarios, including those where the outbreak cannot be contained and the number of cases becomes large. The model accounts for the development of herd immunity by reducing the number of new infections in proportion to the number of susceptible individuals remaining in the population (see Methods). Results of a long-term simulation under scenario B (realistic control effects with slow case isolation, reflecting that this becomes more difficult once case numbers accelerate) are shown in Figure 8 for a total population size of 5 million people.

**Figure 8:**
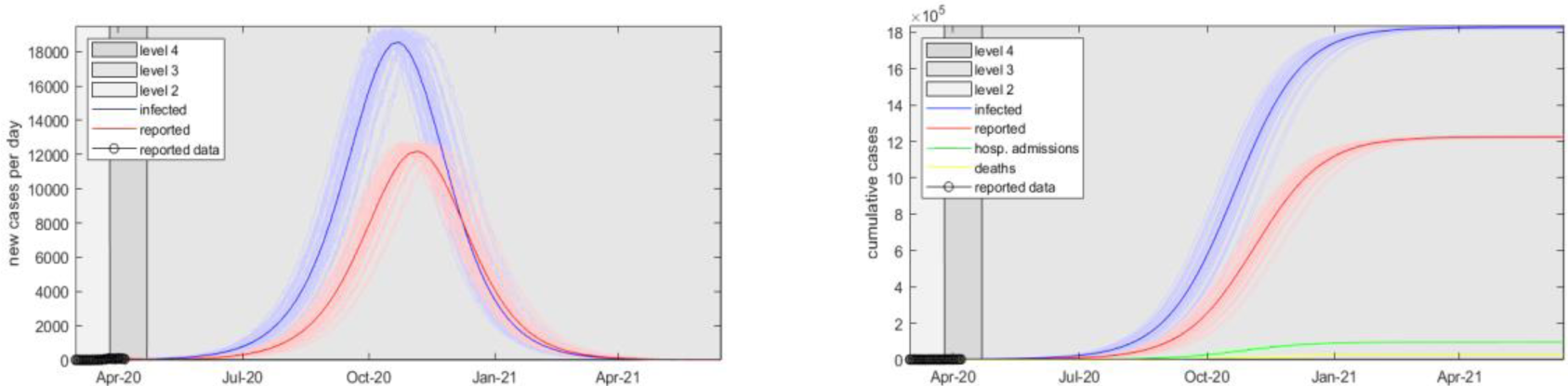
A long-term simulation under scenario B (realistic control efficacy and slow case isolation) with Alert Level 4 control in place for 28 days, followed by Alert Level 3. Deaths are calculated using a baseline infection fatality rate of 0.88% and an elevated rate at times when hospital capacity is exceeded up to a maximum infection fatality rate of 1.65%.

## Discussion

Our results show that for small COVID-19 case numbers, such as those New Zealand is currently managing, rapid case isolation can play an important role. This opens up the possibility of containment or elimination, scenarios that are very distant for most other countries. We summarise the results of the scenarios considered here in Table 3. An optimistic scenario with strong effective controls and rapid case isolation can contain the outbreak at the end of the four week Level 4 period. Other scenarios where the Level 4 controls are relaxed at the end of four weeks lead to a further outbreak. These scenarios would require the re-imposition of Level 4 control at a later stage if this second outbreak were to be controlled. Longer control periods allow for containment in realistic scenarios with rapid case isolation. Longer control periods are more likely to be able to reduce the number of cases so levels where COVID-19 might be eliminated.

The effective reproduction numbers used in the optimistic, realistic, and pessimistic scenarios are comparable to those observed in other countries that have enacted strong population-wide controls. This should give some confidence in the utility of the scenarios considered here, although ideally the control function *C*(*t*) should be determined from an understanding of the underlying contact structure in New Zealand. This will be considered in future work.

We have also shown that rapid case isolation while case numbers are low can drive these effective reproduction numbers even lower. The results also show that an indication of New Zealand’s effective reproduction numbers should emerge in the coming weeks, giving a good sense of the likelihood of containment if numbers fall. Nevertheless, longer periods of strong population-wide controls will delay or reduce the risk of a further outbreak.

Our results also suggest that different testing strategies (e.g. testing of essential services personnel with high numbers of contacts) followed by case isolation and contact tracing should also be considered as an additional control measure. New technologies that allow for faster contact tracing or testing should also be investigated. Improving the speed at which cases are isolated, by testing, tracing, or otherwise, will increase the chance of containment and elimination.

It is important to stress the limitations of the model for assessing long-term impacts. Further versions of this model will include an age-structured population with the possibility that contact rates between age groups or other demographic groupings could be differentially affected by specific control interventions. It will be important to consider the effects of geographic dispersion and contact network structure. This will allow for investigation of regional containment combined with inter-regional travel restrictions, selective reopening of schools and businesses, and potential harms to at risk communities and essential workers.

The branching process model assumes that all infected individuals in either the clinical category or the subclinical category have the same transmission rate. This understates the effect of stochasticity and demographic variability among individuals. In reality, there will be a distribution of transmission rates and the model could be generalised to include this. The effect of this is to increase variance in the model trajectories, and this tends to results in a higher proportion of realisations ending in elimination, but conversely faster growing outbreaks for realisations that do not end in elimination (Lloyd-Smith et al, 2005).

The model can be used to test potential strategies for shifting between alert levels based on, for example, newly reported cases or current hospital demand. Different types of strategy can be examined, for example, a strategy aimed at elimination might require a period with no new cases to go from alert level 4 to 3. A strategy aimed at containment might move the alert level down if the number of new cases falls below some threshold. A strategy aimed at keeping hospitals under capacity could move the alert level up once the number of patients in ICU exceeds some threshold. These strategies could also be examined at a regional level.

## Data Availability

This article does not present new data.

## Acknowledgements

The authors acknowledge the support of Statistics New Zealand, ESR, and the Ministry of Health in supplying data in support of this work. The authors would also like to thank Dr Matt Parry, Professor Nigel French, Dr Anya Mizdrak, Dr Fraser Morgan, Dr Markus Luczak-Roesch, and Dr Samik Datta for their useful comments on the manuscript.

## Footnotes

1 See www.covid19.govt.nz

## References

Binny RN, Hendy, SC, James A, Lustig, A, Plank MJ, Steyn N, (2020. Effect of Level 4 control on R0: review of international COVID-19 cases. Forthcoming.

CDC (2020). Severe Outcomes Among Patients with Coronavirus Disease 2019 (COVID-19) — United States, February 12–March 16, 2020. Morbidity and Mortality Weekly Report. Retrieved from https://www.cdc.gov/mmwr/volumes/69/wr/mm6912e2.htm

Davies NG, Kucharski AJ, Eggo RM, Gimma A, Edmunds WJ (1 April 2020) The effect of non-pharmaceutical interventions on COVID-19 cases, deaths and demand for hospital services in the UK: a modelling study, London School of Hygiene & Tropical Medicine.

Ferguson NM et al & (16 March 2020). Impact of non-pharmaceutical interventions (NPIs) to reduce COVID-19 mortality and healthcare demand. Imperial College COVID-19 Response Team.

Ferretti, L., et al (2020). Quantifying SARS-CoV-2 transmission suggests epidemic control with digital contact tracing. Science. DOI: 10.1126/science.abb6936

Flaxman, S., Swapnil M., and Gandy, A (2020). “Estimating the number of infections and the impact of non-pharmaceutical interventions on COVID-19 in 11 European countries. ” Imperial College COVID-19 Response Team 30.

Ganyani T, Kremer C, Chen D, Torneri A, Faes C, Wallinga J, Hens N (2020). Estimating the generation interval for COVID-19 based on symptom onset data. MedRxiv 2020 (8 March). DOI: 10.1101/2020.03.05.20031815v1.

James A, Hendy SC, Plank MJ, Steyn N (2020). Suppression and mitigation strategies for control of COVID-19 in New Zealand. MedRxiv DOI: 10.1101/2020.03.26.20044677.

Kucharski AJ, Russell TW, Diamond C, Liu Y, Edmunds J, Funk S, and Eggo RM (2020). Early dynamics of transmission and control of COVID-19: a mathematical modelling study. The Lancet. DOI: 10.1016/S1473-3099(20)30144-4

Lauer, S. A., et al (2020). The incubation period of coronavirus disease 2019 (COVID-19) from publicly reported confirmed cases: estimation and application. Annals of internal medicine.

Lloyd-Smith, J. O., Schreiber, S. J., Kopp, P. E., & Getz, W. M. (2005). Superspreading and the effect of individual variation on disease emergence. Nature, 438(7066), 355–359.

Ministry of Health (2005). Intensive Care Services in New Zealand: A report to the Deputy Director-General, Clinical Services. Wellington: Ministry of Health.

Ministry of Health (2020). COVID-19 (novel coronavirus). Current Situation 1 April 2020.

StatsNZ. (n.d.). Infoshare. Retrieved Jan 19, 2020, from http://archive.stats.govt.nz/infoshare/

Verity R et al (2020). Estimates of the severity of coronavirus disease 2019: a model-based analysis. The Lancet https://doi.org/10.1016/S1473-3099(20)30243-7

World Health Organisation (28 February 2020). Report of the WHO-China Joint Mission on Coronavirus Disease 2019 (COVID-19)

World Health Organisation (11 March 2020). Coronavirus disease 2019 (COVID-19) Situation Report – 51.

Wilson N, Telfar Barnard L, Kvalsig A, Verrall A, Baker MG, Schwehm M. (2020) Modelling the Potential Health Impact of the COVID-19 Pandemic on a Hypothetical European Country. medRxiv 2020.03.20.20039776

Zhou F, Yu T, Du R, Fan G, Liu Y, Liu Z, Xiang J, Wang Y, Song B, Gu X, Guan L, Wei Y, Li H, Wu X, Xu J, Tu S, Zhang Y, Chen H, Cao B (2020). Clinical course and risk factors for mortality of adult inpatients with COVID-19 in Wuhan, China: a retrospective cohort study. Lancet 395: 1054–1062. doi: 10.1016/S0140-6736(20)30566-3

